# Dynamics of anti-Spike IgG antibody level after full BNT162b2 COVID-19 vaccination in health care workers

**DOI:** 10.1101/2021.12.14.21267783

**Authors:** Hiroaki Ikezaki, Ryoko Nakashima, Kahori Miyoshi, Yuichi Hara, Jun Hayashi, Hiroshi Hara, Hideyuki Nomura, Nobuyuki Shimono

**Author notes:** **Corresponding author:** Hiroaki Ikezaki, MD. PhD., Associate Professor., Department of Comprehensive General Internal Medicine, Kyushu University Faculty of Medical Sciences., 3-1-1 Maidashi, Higashi-Ku, Fukuoka, 812-8582, Japan. **Contribution statement:** The conception and design of the study was carried out by HI, HN, and NS. The acquisition of data was done by all authors. The measurement was done by HI. The data were analyzed by HI and interpreted by all co-authors. All authors contributed to the drafting of the paper and its revision and are responsible for the intellectual content and the final approval of the version to be published. HI is the guarantor of this study.

## Abstract

**Introduction:** The administration of a third vaccine is ongoing in many countries, but the evaluation of vaccine-induced immunity is still insufficient. This study evaluated anti-spike IgG levels in 373 health care workers six months after the BNT162b2 vaccination.

**Methods:** Dynamics of anti-spike IgG levels six months after the 2nd vaccination were assessed in 49 participants (Analysis-1). A cross-sectional assessment of anti-spike IgG level was performed in 373 participants (Analysis-2). Participants positive for anti-nucleocapsid IgG or IgM and receiving immunosuppressants were excluded from Analysis-2.

**Results:** In Analysis 1, the median anti-spike IgG level was lower in the older age group and decreased consistently after the second vaccination regardless of age. In Analysis-2, the anti-spike IgG level was significantly negatively associated with age (r = −0.35, p < 0.01). This correlation remained statistically significant (r = −0.28, p < 0.01) even after adjusting for sex, BMI, smoking habits, alcohol drinking habits, allergies, and the presence of fever or other adverse reactions at the time of the vaccination. Alcohol drinking habit was also associated with the anti-spike IgG level; daily alcohol drinkers had significantly lower anti-spike IgG levels than never alcohol drinkers. Sex, smoking habit, allergy, and fever and other side effects after the vaccination were not associated with anti-spike IgG levels six months after the 2nd vaccination.

**Conclusions:** Six months after the vaccination, the anti-spike IgG level was substantially low among older persons and daily alcohol drinkers.

## Introduction

The mRNA coronavirus disease 2019 (COVID-19) vaccines, BNT162b2 (Pfizer/BioNTech) and mRNA-1273 (Moderna), have shown very high effectiveness in preventing COVID-19 in real-world practice [1, 2]. However, many countries are currently experiencing a resurgence of COVID-19, dominated by the Delta (B.1.617.2) variant of SARS-CoV-2 [3, 4]. In addition, a new variant of SARS-CoV-2, B.1.1.529, was reported, and the World Health Organization named this new variant Omicron and classified it as a Variant of Concern on November 26, 2021 [5]. Because the Omicron strain is more transmissible than previous strains, including delta strains, and can reinfect people who have already had the virus or been vaccinated, concerns about a resurgence of the pandemic are rising [6, 7]. Japan has experienced its fifth pandemic wave since late June 2021, with up to 25,000 infections per day (20 / 100,000) and more than 800,000 hospitalizations in total (650 / 100,000) [8]. However, it is difficult to estimate the vaccine’s effectiveness in the Japanese population, as the vaccination rate during the peak of the fifth pandemic wave in Japan was only about 40% [9]. The increase in infections and hospitalizations of vaccinated individuals in other countries with high vaccination rates potentially stems from a combination of waning vaccine immunity over time and potentially reduced vaccine effectiveness against the delta variant [10-13]. In response to the resurgence of COVID-19, many countries are starting the administration of a third vaccination, and Israel was the first country to start the third vaccination in July 2021 [14, 15]. The decrease in immunoglobulin G (IgG) level to spike protein six months after the second vaccination has been cited as a rationale for the third vaccination. However, it is still unclear how much IgG level is enough to prevent COVID-19 infection. In addition, although the vaccine’s excellent effectiveness in preventing symptomatic infection in the early stage after the third vaccination has been reported, it is also unclear how long the effect will be maintained [14, 15]. On the other hand, there are some reports that the effect of preventing severe disease is maintained even six months after the second vaccination [11, 16]. However, it is also unclear how much IgG level is enough to avert COVID-19 severe disease. Therefore, it is challenging to determine target individuals for the additional vaccination.

This study aims to investigate the dynamics of the anti-spike IgG levels in the health care workers during six months after being fully vaccinated and find indicators to whom we should strongly recommend the additional vaccination.

## Methods

### Study participants

Study participants were recruited from health care workers in Haradoi Hospital, a care-mix hospital in Fukuoka. Of the total 485 health care workers in this hospital, 373 (76.9%) participated in this study. Because most of the study participants were nurses, approximately 80% of the participants were women. All participants provided written informed consent prior to enrollment. All studies were carried out in accordance with the principles of the Declaration of Helsinki, as revised in 2008, and approved by the Haradoi hospital institutional ethics review committee prior to data collection (Approval No. 2020-08).

This study consisted of two sub-studies: the first was to evaluate the dynamics of anti-spike IgG levels by measuring IgG antibody levels before the first vaccination, three weeks after the first vaccination (just before the second vaccination), and one month, two months, four months, and six months after the second vaccination; included 49 participants in the first sub-study (Analysis-1). The second was to evaluate IgG antibody levels six months after the second vaccination, and 373 participants, including above 49 Analysis-1 participants, participated in the second sub-study (Analysis-2). All participants were offered first and second doses of vaccine between March and April 2021. Participants also provided information on their height, weight, smoking habits (current, past, or never), drinking habits (daily, often, or never), allergies, medical history, medications, and whether they had adverse reactions to vaccination (fever and other adverse reactions) and antipyretics. We performed additional IgG / IgM antibody qualitative tests against SARS-CoV-2 nucleocapsid protein for participants whose anti-spike IgG antibodies were in the top 10% (2100 AU/ml) in Analysis-2 participants. Of the 373, we excluded the following five participants from the Analysis-2. One was on corticosteroid treatment. Another was infected COVID-19, confirmed by PCR test approximately one month before blood collection. The other three were positive for IgG / IgM antibody against nucleocapsid protein. Therefore, the Analysis-2 was performed on 368 subjects (**Figure 1**).

**Figure 1.**
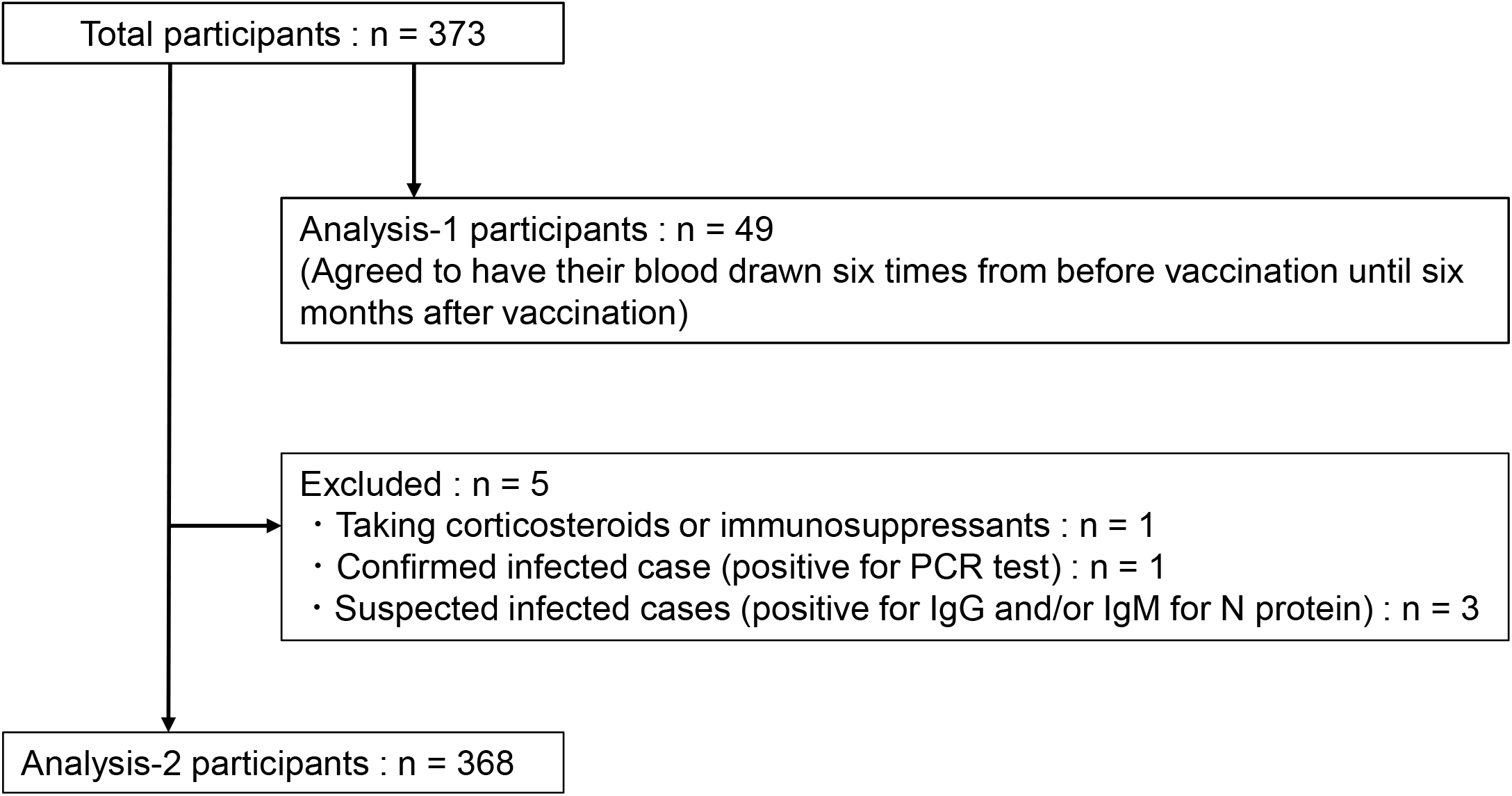
Study population. This study consisted of two sub-studies. Participants in Analysis-1 are also included in Analysis-2.

### Laboratory measurement

Levels of anti-spike IgG were quantified using the SARS-CoV-2 IgG II Quant assay (Abbott Diagnostics, Chicago, IL, USA). Participants in Analysis-1 underwent blood testing to quantitatively assess anti-spike IgG six times (before the first vaccination, just before the second vaccination, and one month, two months, four months, and six months after the second vaccination). All participants in Analysis-2 underwent blood testing to quantitatively assess anti-spike IgG in October 2021, six months after the second vaccination. The results of anti-spike IgG are expressed as arbitrary units per milliliter (AU/ml) (positive threshold: 50 AU/ml; upper limit: 40,000 AU/ml). Additional qualitative testing for IgG and IgM antibodies against nucleocapsid protein was performed using the SARS-CoV-2 IgG and IgM assays (Abbott Diagnostics). We performed these two qualitative tests on participants with anti-spike IgG levels in the top 10% of Analysis-2 participants to assess breakthrough infection. The results of anti-nucleocapsid IgG and IgM antibodies are expressed as index (S/C) (positive thresholds: 1.40 index [S/C] for IgG and 1.00 index [S/C] for IgM). Participants with positive anti-nucleocapsid IgG or IgM were excluded from Analysis-2. Participants in Analysis-1 were also given blood tests for total bilirubin, aspartate aminotransferase (AST), alanine aminotransferase (ALT), γ-glutamyl transpeptidase (γ-GTP), and serum creatinine, using standard enzymatic methods.

### Statistical analysis

All analyses were performed using SAS version 7.4 (SAS Institute Inc). Data are expressed as median values with 25^th^ and 75^th^ percentile values for continuous variables. Categorical variables are reported as frequencies and percentages. The Mann-Whitney U test was used to compare two groups, and the Kruskal-Wallis test was used to compare three or more groups. The Tukey-Kramer method was used for each two-group comparison among three or more groups. Anti-spike IgG level with adjustment factors was determined by the least means square method. Univariate analysis for categorical variables was carried out using the Chi-square test or Fisher’s exact test. Odds ratios for each factor for anti-spike IgG levels of ≥1000 or ≥2150 AU/ml at six months were determined by univariate and multivariate logistic analyses. In the multivariate logistic regression analysis, adjustment factors were sex, age, smoking habits, alcohol drinking habits, allergy, and the interval days between the second vaccination and anti-spike IgG measurement. The statistical significance threshold was set at 5%.

## Results

### Analysis-1

**Table 1** shows baseline characteristics of 49 participants of Analysis-1. Of those analyzed, the median age was 41 years, the median BMI was 21.2 kg/m^2^, 85.7% were women, and all women were under 60 years of age. There was no current smoker, 10.2% were daily alcohol drinkers, and 18.4% had an allergy. The anti-spike IgG level of all 49 participants before the first vaccination was below 50 AU/ml. In other blood tests, AST, ALT, γ-GTP, and serum creatinine levels were higher in the group over 60 years than in the two younger groups. There was no significant difference in having fever and other adverse reactions after the vaccinations between the age groups. The prevalence of having antipyretics was lower in the group over 60 years than in the two younger groups.

**Table 1.** Baseline characteristics of participants in Analysis 1 ^†^

|  | 20s and 30s (n = 22) | 40s and 50s (n = 23) | 60s and older (n = 4) |
| --- | --- | --- | --- |
| **Demographic** |  |  |  |
| Age – years | 30 [27, 35] | 46 [42, 50] | 69 [66.5, 79.5] |
| Sex – no. (%) |  |  |  |
| Female | 21 (95.5) | 21 (91.3) | 0 (0.0) |
| Male | 1 (4.5) | 2 (8.7) | 4 (100.0) |
| Body mass index<sup>‡</sup> – kg/m<sup>2</sup> | 20.2 [18.6, 21.9] | 22.9 [20.4, 25.7] | 22.5 [19.0, 23.3] |
| Smoking habit – no. (current / past / never) | 0 / 2 / 20 | 0 / 3 / 20 | 0 / 1 / 3 |
| Alcohol drinking habit – no. (daily / often / never) | 1 / 13 / 8 | 3 / 15 / 5 | 1 / 1 / 2 |
| Allergy – no. (%) | 6 (27.3) | 3 (13.0) | 0 (0.0) |
| **Comorbidities** |  |  |  |
| Number of comorbidities – no. | 0 [0, 1] | 0 [0, 1] | 2 [1, 4] |
| Hypertension – no. (%) | 1 (4.5) | 3 (13.0) | 2 (50.0) |
| Diabetes – no. (%) | 2 (9.1) | 0 (0.0) | 2 (50.0) |
| Dyslipidemia – no. (%) | 1 (4.5) | 3 (13.0) | 2 (50.0) |
| Hyperuricemia – no. (%) | 1 (4.5) | 0 (0.0) | 1 (25.0) |
| Coronary heart disease – no. (%) | 0 (0.0) | 0 (0.0) | 0 (0.0) |
| Arrhythmia – no. (%) | 1 (4.5) | 1 (4.4) | 1 (25.0) |
| Stroke – no. (%) | 0 (0.0) | 0 (0.0) | 0 (0.0) |
| Lung diseases – no. (%) | 0 (0.0) | 2 (8.7) | 0 (0.0) |
| Thyroid disease – no. (%) | 1 (4.5) | 0 (0.0) | 0 (0.0) |
| Atopic dermatitis – no. (%) | 2 (9.1) | 1 (4.4) | 0 (0.0) |
| Autoimmune diseases – no. (%) | 0 (0.0) | 0 (0.0) | 0 (0.0) |
| Cancer – no. (%) | 0 (0.0) | 0 (0.0) | 1 (25.0) |
| **Laboratory measurement** |  |  |  |
| Total bilirubin – mg/dl | 0.6 [0.5, 0.9] | 0.5 [0.5, 0.7] | 0.8 [0.65, 0.95] |
| Aspartate aminotransferase – IU/l | 18 [15, 21] | 19.5 [15, 23] | 25.5 [23.5, 29.5] |
| Alanine aminotransferase – IU/l | 14 [9, 18] | 15 [12, 19] | 21 [19, 24] |
| γ-glutamyl transpeptidase – IU/l | 15 [12, 19] | 17.5 [15, 35] | 23.5 [22, 28] |
| Serum creatinine – mg/dl | 0.59 [0.53, 0.63] | 0.64 [0.58, 0.70] | 0.83 [0.76, 0.91] |
| **Side effect of vaccination** |  |  |  |
| Fever – no. (%) | 12 (54.6) | 13 (56.5) | 2 (50.0) |
| Other side effect – no. (%) | 19 (86.4) | 18 (78.3) | 3 (75.0) |
| Antipyretics – no. (%) | 16 (72.7) | 18 (78.3) | 1 (25.0) |

<sup>†</sup> Continuous variables are presented as median [1<sup>st</sup> quartile, 3<sup>rd</sup> quartile], and categorical variables are presented as number (%).
<sup>‡</sup> Body mass index was calculated using the following equation: body weight (kg) / height (m) / height (m).

Dynamics in anti-spike IgG level by age group is shown in **Figure 2**. After the first vaccination, all 49 participants had an IgG level ≥ 50 AU/ml. The median anti-spike IgG levels after the first vaccination were higher in the younger age group than the older age group: 1680.5 AU/ml in the 20s and 30s group, 1049.2 AU/ml in the 40s and 50s group, and 151.2 AU/ml in the group over 60 years, respectively. This trend continued after the second vaccination. In this analysis, the anti-spike IgG level peaked one month after the second vaccination: 14000 AU/ml in the 20s and 30s group, 9835.3 AU/ml in the 40s and 50s group, and 4538.4 AU/ml in the group over 60 years, respectively. There was no difference in the anti-spike IgG decline rate from one month to six months after the second vaccination: 91.9% in the 20s and 30s group, 93.1% in the 40s and 50s group, and 93.2% in the group of over 60 years, respectively. The median anti-spike IgG levels at six months after the second vaccination were 1033.7 AU/ml in the 20s and 30s group, 710.4 AU/ml in the 40s and 50s group, and 464.6 AU/ml in the group over 60 years, respectively.

**Figure 2.**
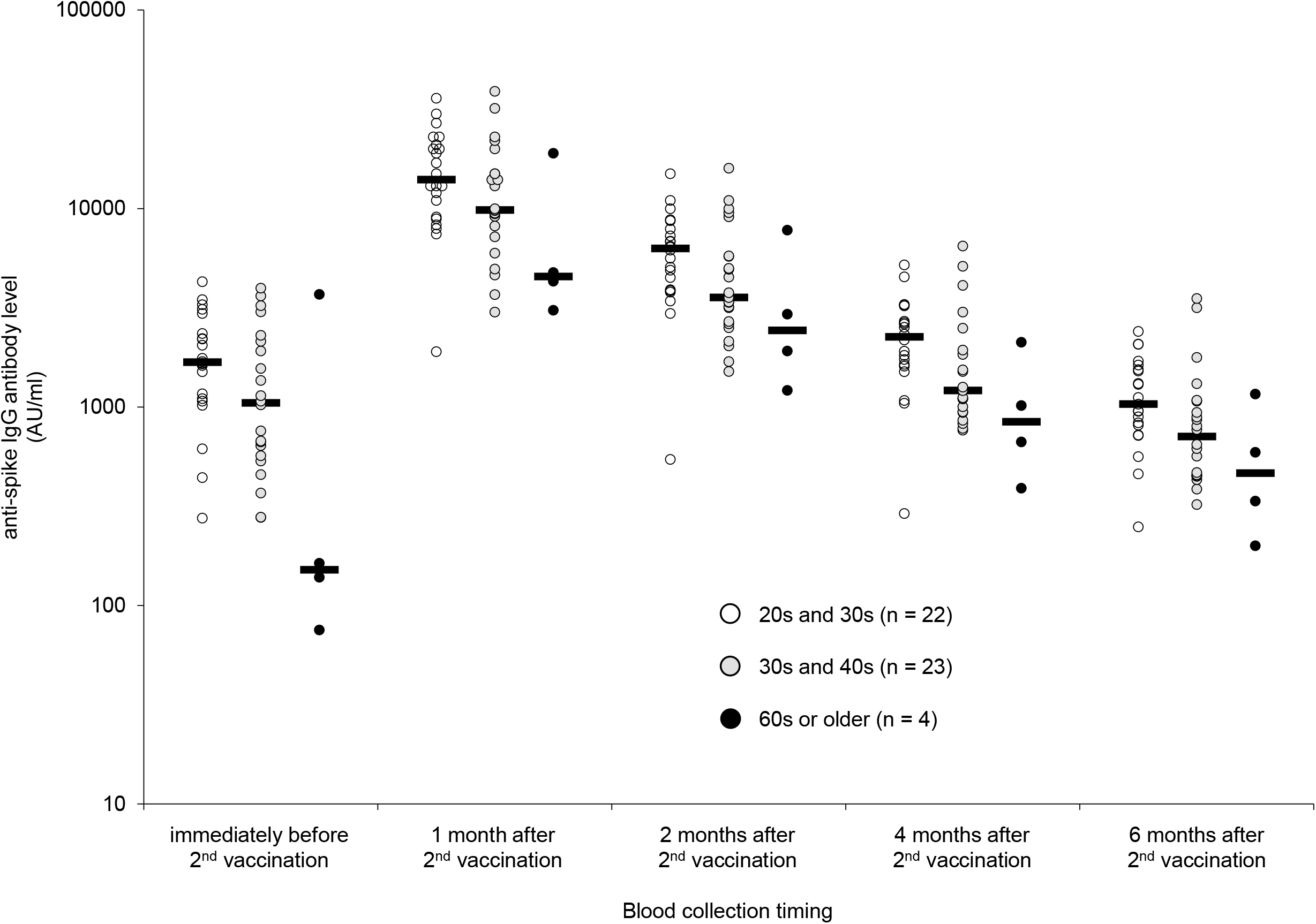
Distribution of anti-spike IgG levels after the 2^nd^ vaccination. Distribution of anti-spike IgG levels by age group is shown. The anti-spike IgG levels were measured three weeks after the first vaccination (just before the second vaccination) and one month, two months, four months, and six months after the second vaccination. Dots represent individual observed serum samples. Black bars indicate median anti-spike IgG levels.

### Analysis-2

Baseline characteristics of 373 participants of Analysis-2 is shown in **Table 2**. The demographic of participants was similar to Analysis-1 participants: median age was 42 years, median BMI was 21.3 kg/m^2^, and 79.9% were women. Regarding the side effect of vaccination, 48.0% experienced fever, 66.5% experienced other side effects, and 55.5% took antipyretics. Of the Analysis-2 participants, 5.9% were current smokers, 14.2% were daily alcohol drinkers, and 13.1% had an allergy. The median interval between the date of the second vaccination and the date of blood collection was 185 days. One of the subjects took immunosuppressive drugs, including steroids, and was excluded from the Analysis-2 (**Table 2 and Figure 1**). In addition, we performed testing for anti-nucleocapsid IgM and IgG antibodies on participants with anti-spike IgG levels in the top 10% of Analysis-2 participants. Four participants with positive anti-nucleocapsid IgM or IgG were excluded from Analysis-2 because of possible COVID-19 breakthrough infection (**Figure 1**).

**Table 2.**
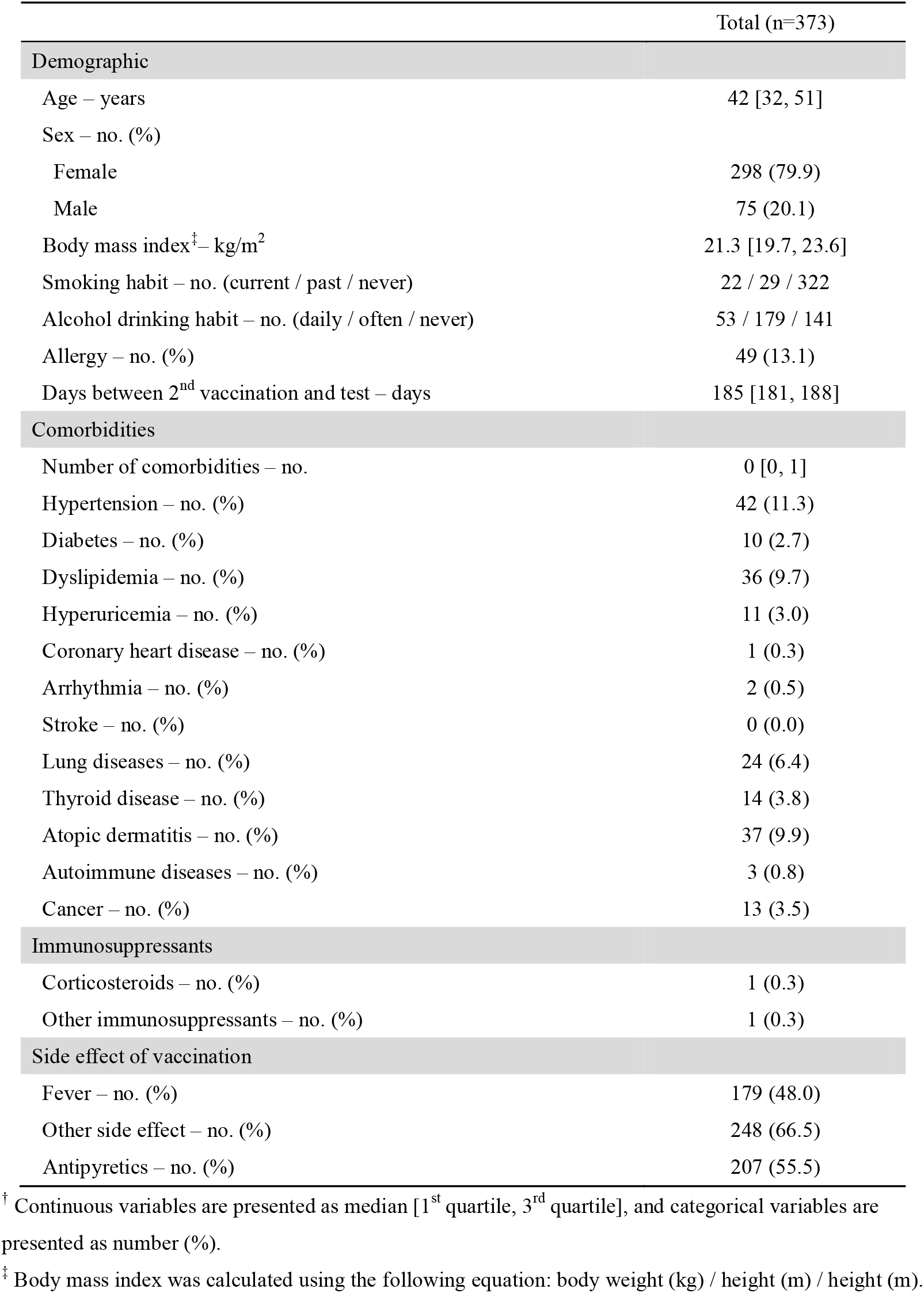
Baseline characteristics of participants in Analysis 2 ^†^

|  | Total (n=373) |
| --- | --- |
| **Demographic** |  |
| Age – years | 42 [32, 51] |
| Sex – no. (%) |  |
| Female | 298 (79.9) |
| Male | 75 (20.1) |
| Body mass index<sup>‡</sup> – kg/m<sup>2</sup> | 21.3 [19.7, 23.6] |
| Smoking habit – no. (current / past / never) | 22 / 29 / 322 |
| Alcohol drinking habit – no. (daily / often / never) | 53 / 179 / 141 |
| Allergy – no. (%) | 49 (13.1) |
| Days between 2<sup>nd</sup> vaccination and test – days | 185 [181, 188] |
| **Comorbidities** |  |
| Number of comorbidities – no. | 0 [0, 1] |
| Hypertension – no. (%) | 42 (11.3) |
| Diabetes – no. (%) | 10 (2.7) |
| Dyslipidemia – no. (%) | 36 (9.7) |
| Hyperuricemia – no. (%) | 11 (3.0) |
| Coronary heart disease – no. (%) | 1 (0.3) |
| Arrhythmia – no. (%) | 2 (0.5) |
| Stroke – no. (%) | 0 (0.0) |
| Lung diseases – no. (%) | 24 (6.4) |
| Thyroid disease – no. (%) | 14 (3.8) |
| Atopic dermatitis – no. (%) | 37 (9.9) |
| Autoimmune diseases – no. (%) | 3 (0.8) |
| Cancer – no. (%) | 13 (3.5) |
| **Immunosuppressants** |  |
| Corticosteroids – no. (%) | 1 (0.3) |
| Other immunosuppressants – no. (%) | 1 (0.3) |
| **Side effect of vaccination** |  |
| Fever – no. (%) | 179 (48.0) |
| Other side effect – no. (%) | 248 (66.5) |
| Antipyretics – no. (%) | 207 (55.5) |

<sup>†</sup> Continuous variables are presented as median [1<sup>st</sup> quartile, 3<sup>rd</sup> quartile], and categorical variables are presented as number (%).
<sup>‡</sup> Body mass index was calculated using the following equation: body weight (kg) / height (m) / height (m).

**Table 3** shows the characteristics of the excluded four participants with positive anti-nucleocapsid IgM or IgG. One was a confirmed COVID-19 case with PCR test approximately one month before the antibody testing and was positive for both anti-nucleocapsid IgM and IgG. Of the other three subjects, two were positive for both anti-nucleocapsid IgM and IgG, and the other one was positive for only anti-nucleocapsid IgG. The confirmed case had a fever but only mild symptoms and did not require hospitalization. The three suspected cases were asymptomatic or had very mild illness, and did not undergo PCR testing. One confirmed and one suspected case of COVID-19 infection belonged to the same department, while the other two suspected cases belonged to different departments. The IgG antibody levels in the departments these four cases belonged to were not different from those in other departments (data are not shown).

**Table 3.** Characteristics of confirmed and suspected infected cases

|  | Age | Sex | Anti-spike IgG level<sup>‡</sup><br>(AU/ml) | Anti-nucleocapsid IgG level<sup>§</sup><br>(Index [S/C]) | Anti-nucleocapsid IgM level<sup>□</sup><br>(Index [S/C]) | Subjective symptoms<sup>¶</sup> |
| --- | --- | --- | --- | --- | --- | --- |
| Confirmed case<sup>†</sup> | 40s | Female | 50,000 | 5.87 | 1.67 | fever, nasal mucus |
| Suspected case 1 | 20s | Female | 70,000 | 5.80 | 14.45 | none |
| Suspected case 2 | 40s | Female | 28,000 | 1.88 | < 1.00 | nasal mucus |
| Suspected case 3 | 50s | Female | 7,029.1 | 1.93 | 1.26 | none |

<sup>†</sup> COVID-19 infection was confirmed by PCR test approximately one month before the blood test.
<sup>‡</sup> Positive threshold is 50 AU/ml.
<sup>§</sup> Positive threshold is 1.40 index (S/C).
<sup>□</sup> Positive threshold is 1.00 index (S/C).
<sup>¶</sup> Subjective symptoms in the confirmed case are at the time of PCR testing and those in suspected cases are during two months prior the blood test.
AU, arbitrary units; IgG, immunoglobulin G.

**Figure 3** shows the anti-spike IgG level distribution at six months after the second vaccination of 368 subjects in Analysis 2 according to age. The anti-spike IgG level showed a statistically significant negative correlation with age (*r* = −0.35, *p* < 0.01). This correlation remained statistically significant (*r* = −0.28, *p* < 0.01) even after adjusting for sex, BMI, smoking habits, alcohol drinking habits, allergies, and the presence of fever or other adverse reactions at the time of the vaccination. Anti-spike IgG levels for various groups are shown in **Figure 4**. The anti-spike IgG levels were adjusted for age (not in **Figure 4a**), sex (not in **Figure 4b**), and the interval days between the second vaccination and anti-spike IgG measurements. The 20s or younger group had significantly higher anti-spike IgG levels than any other group. The 30s group had significantly higher anti-spike IgG levels than the 40s and 60s groups (**Figure 4a**). The current smoker group tended to have lower anti-spike IgG levels than the past and never smoker groups, but the difference was not statistically significant (**Figure 4c**). The group of daily alcohol drinking had significantly lower anti-spike IgG levels than the groups of never alcohol drinking or occasionally alcohol drinking (**Figure 4d**). Other factors such as sex, allergies, and presence of fever or other adverse reactions at the vaccination had no considerable effect on anti-spike IgG level (**Figure 4b, 4e, 4f, and 4g**).

**Figure 3.**
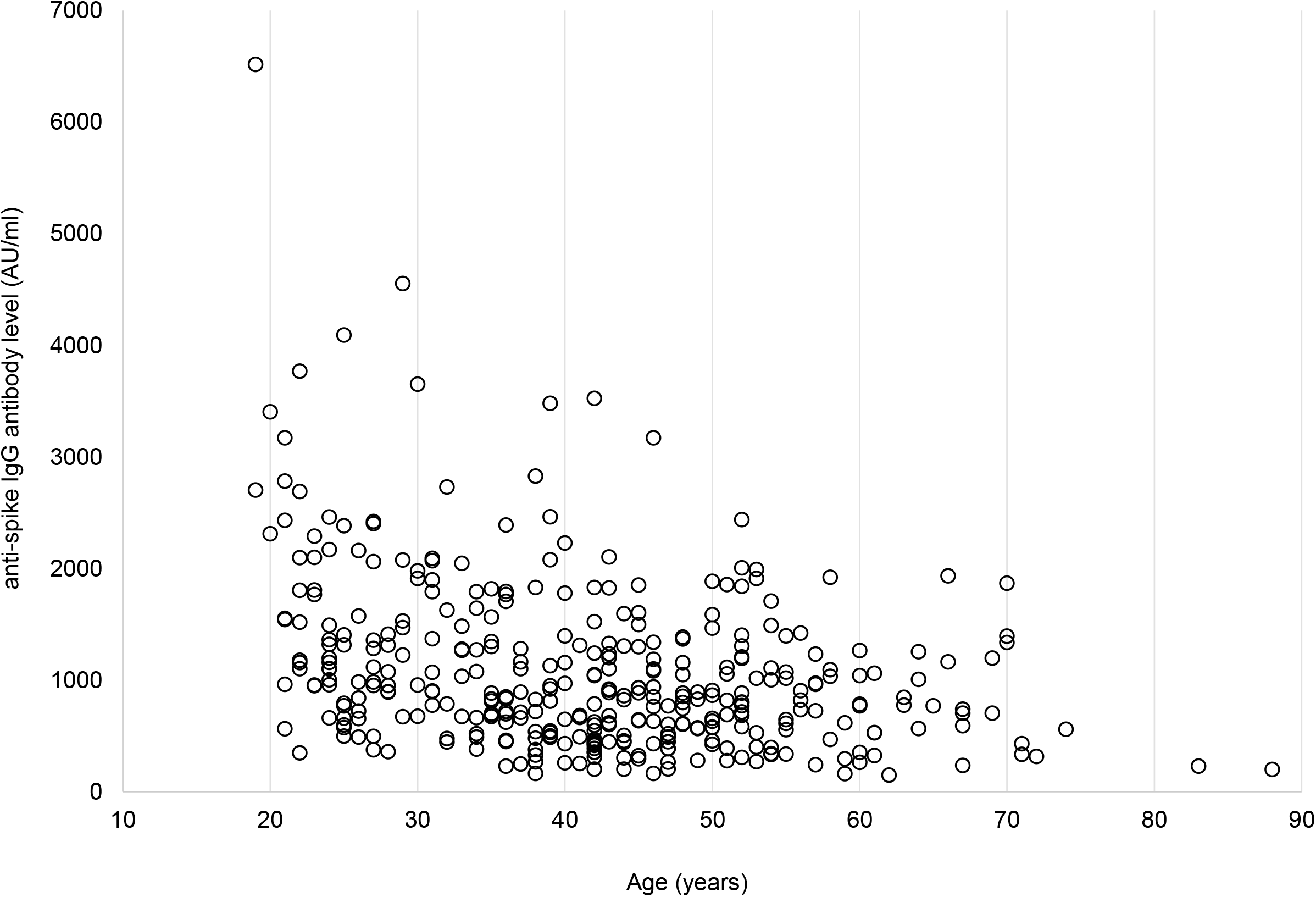
Correlation between age and anti-spike IgG level six months after the 2^nd^ vaccination. Distribution of anti-spike IgG level by age is shown. Dots represent individual observed serum samples.

**Figure 4.**
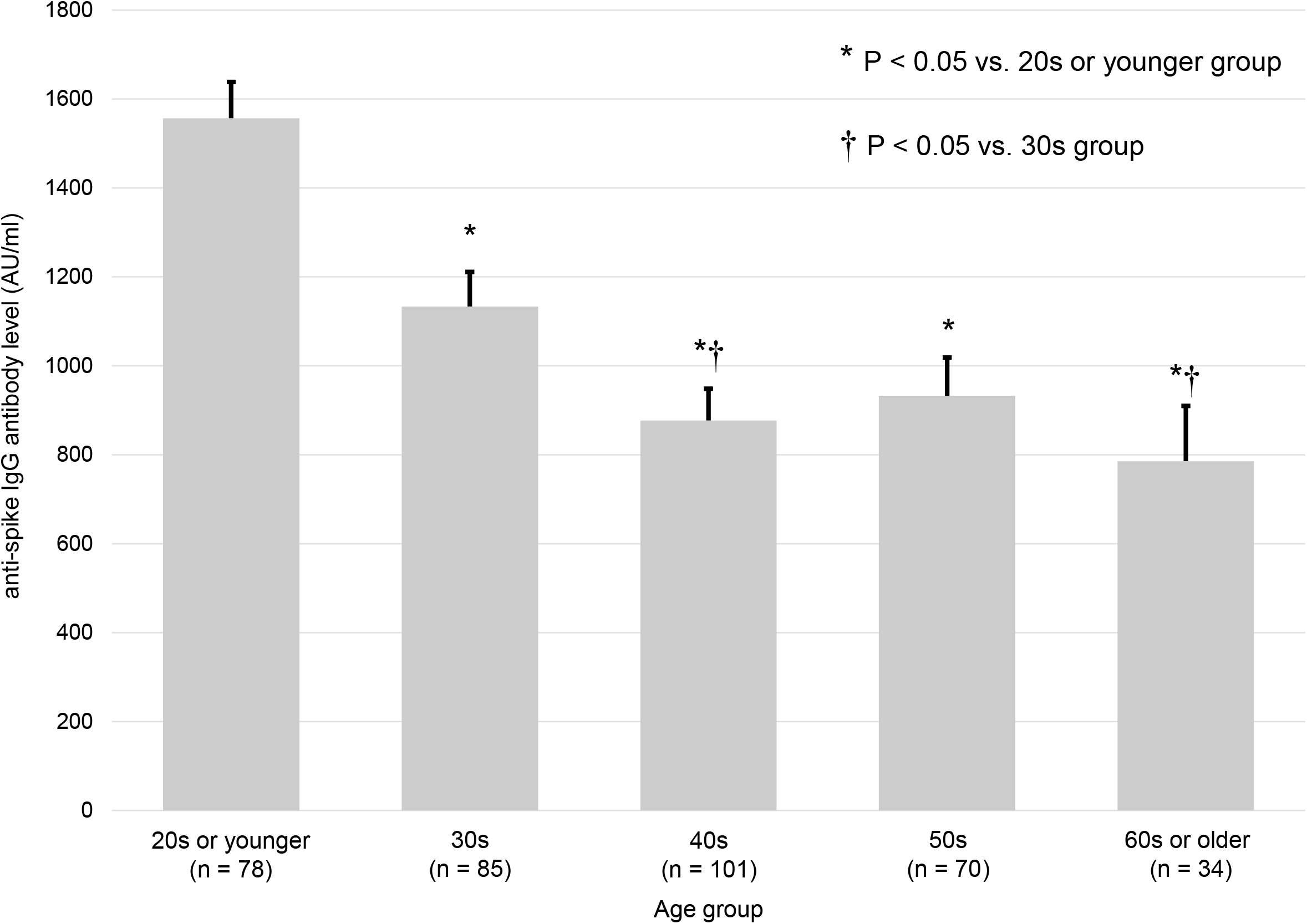
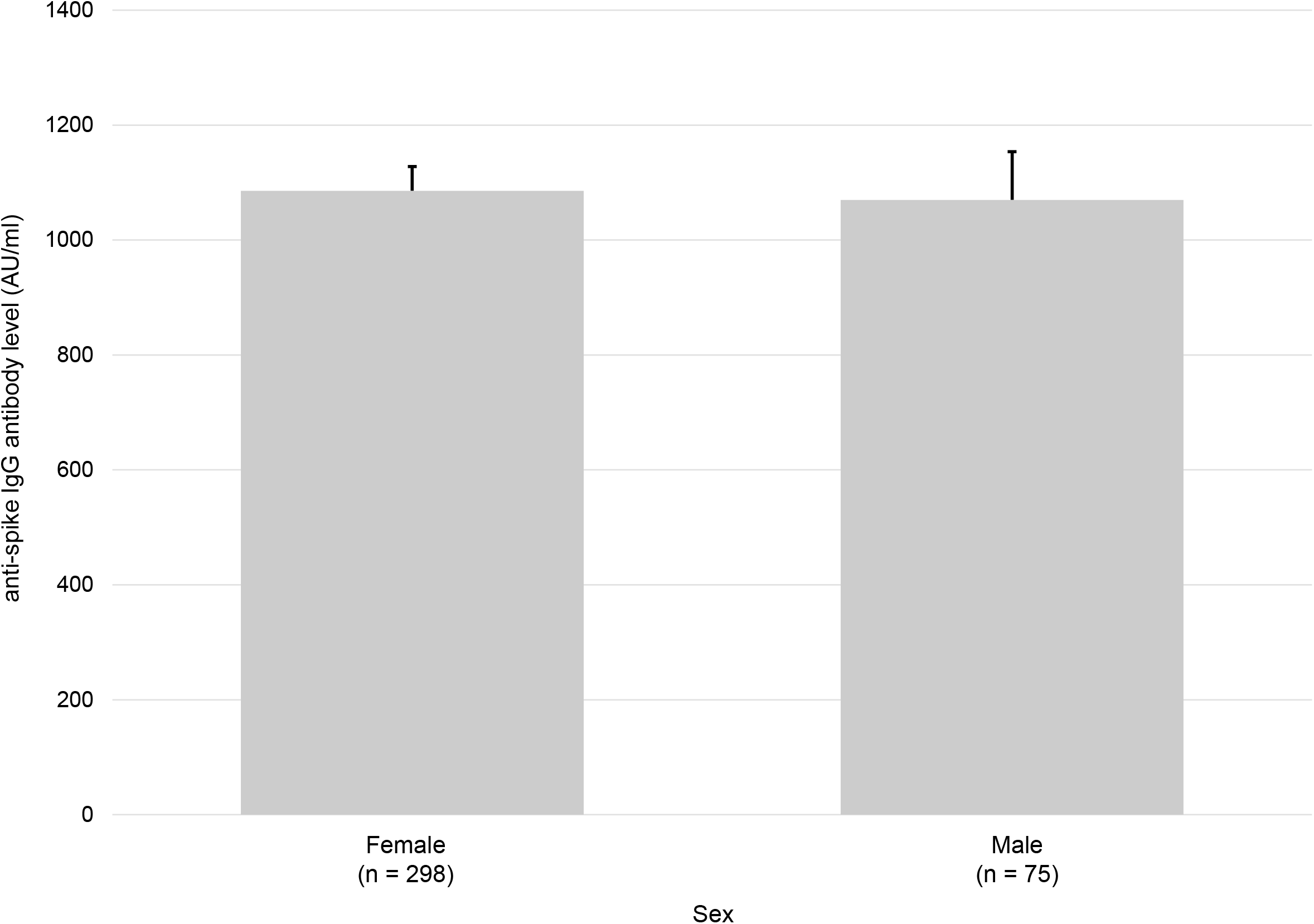
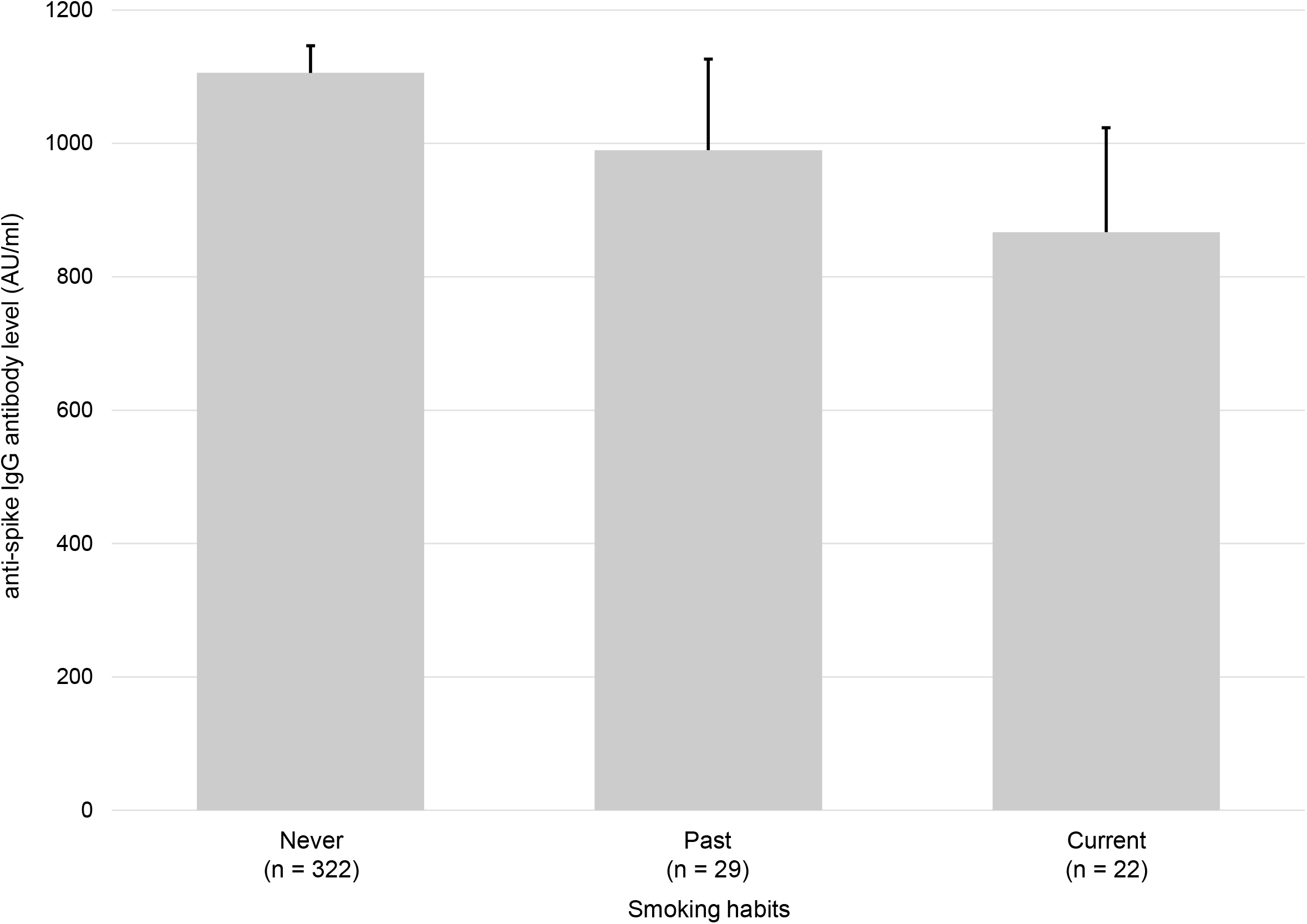
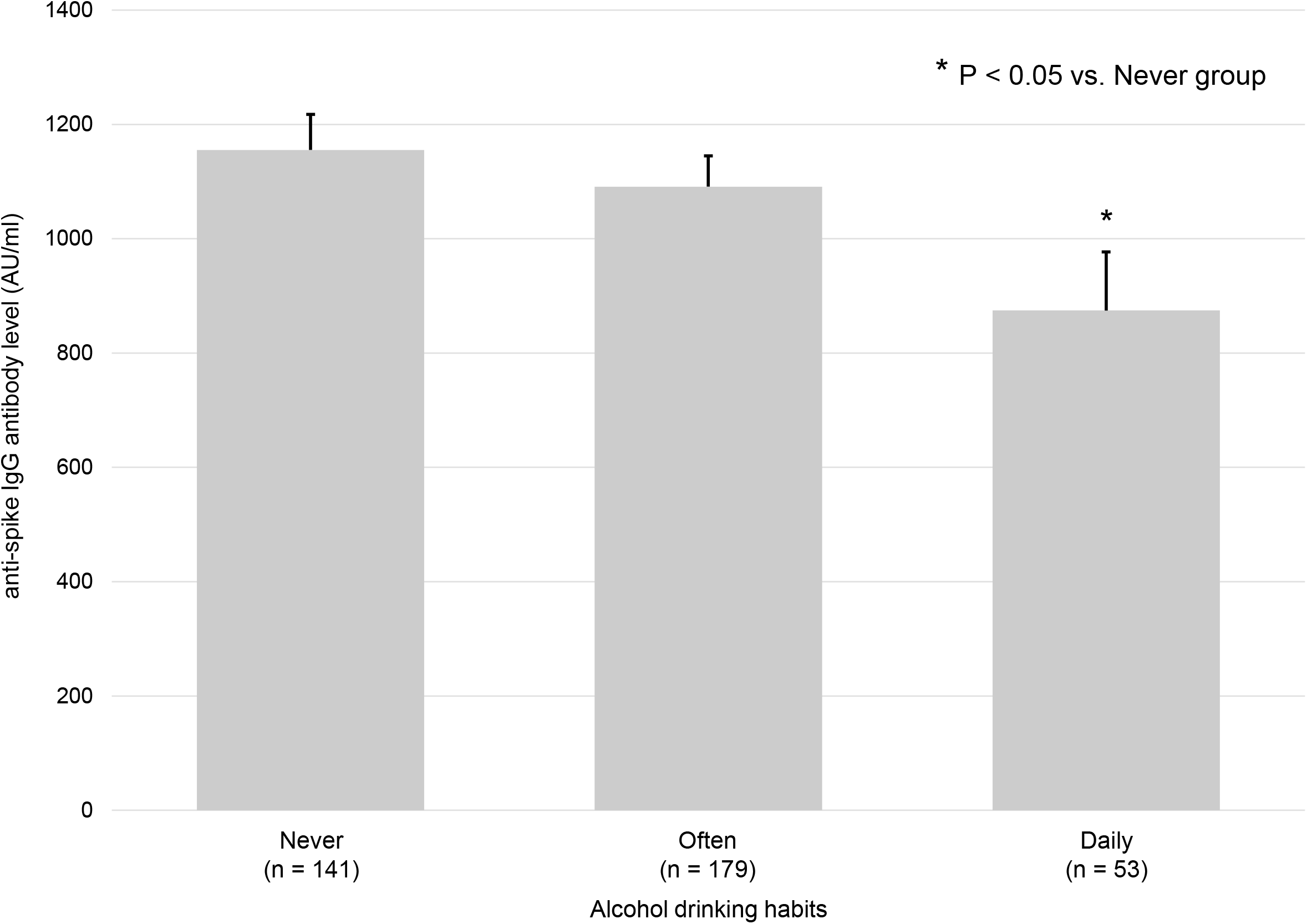
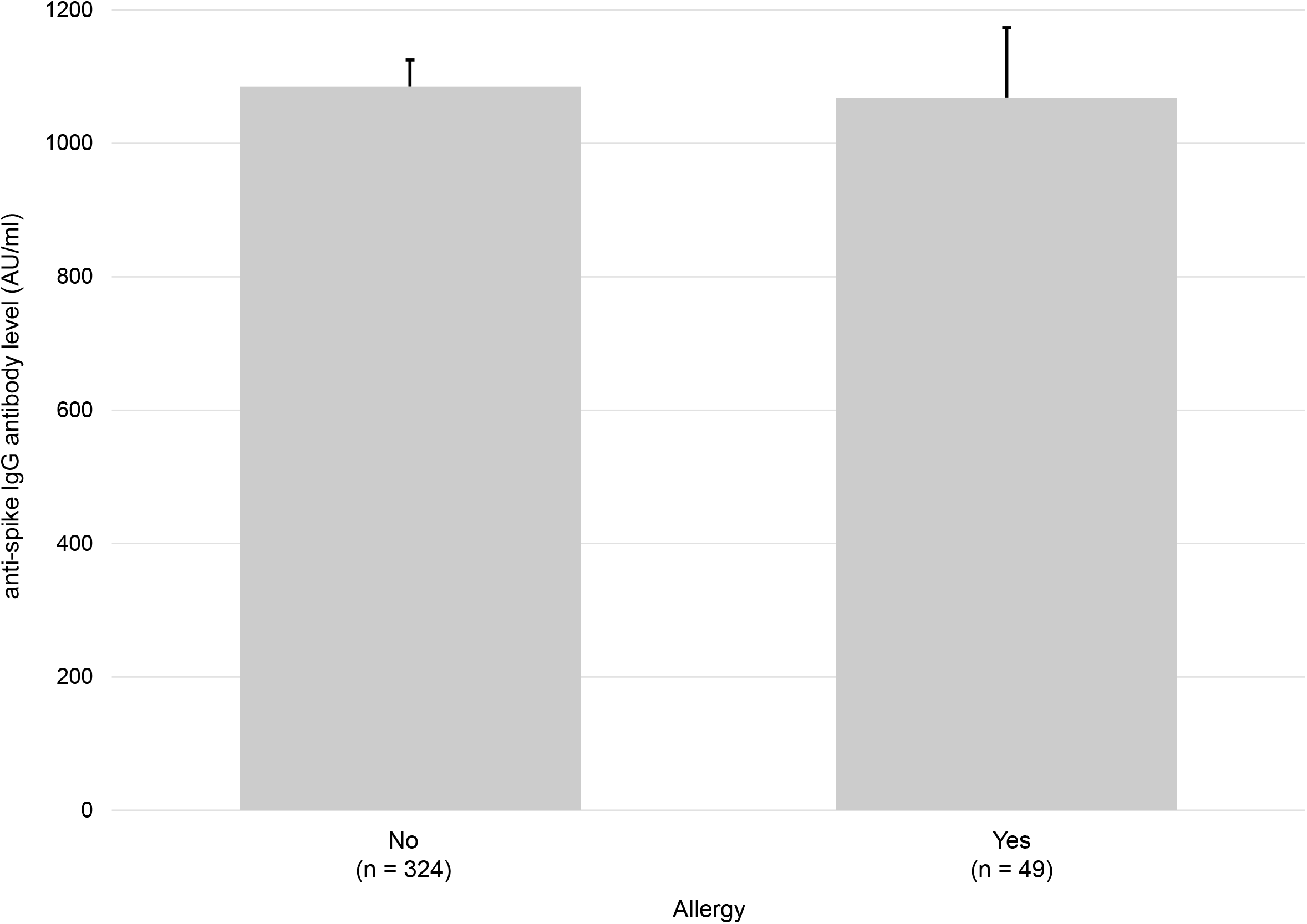
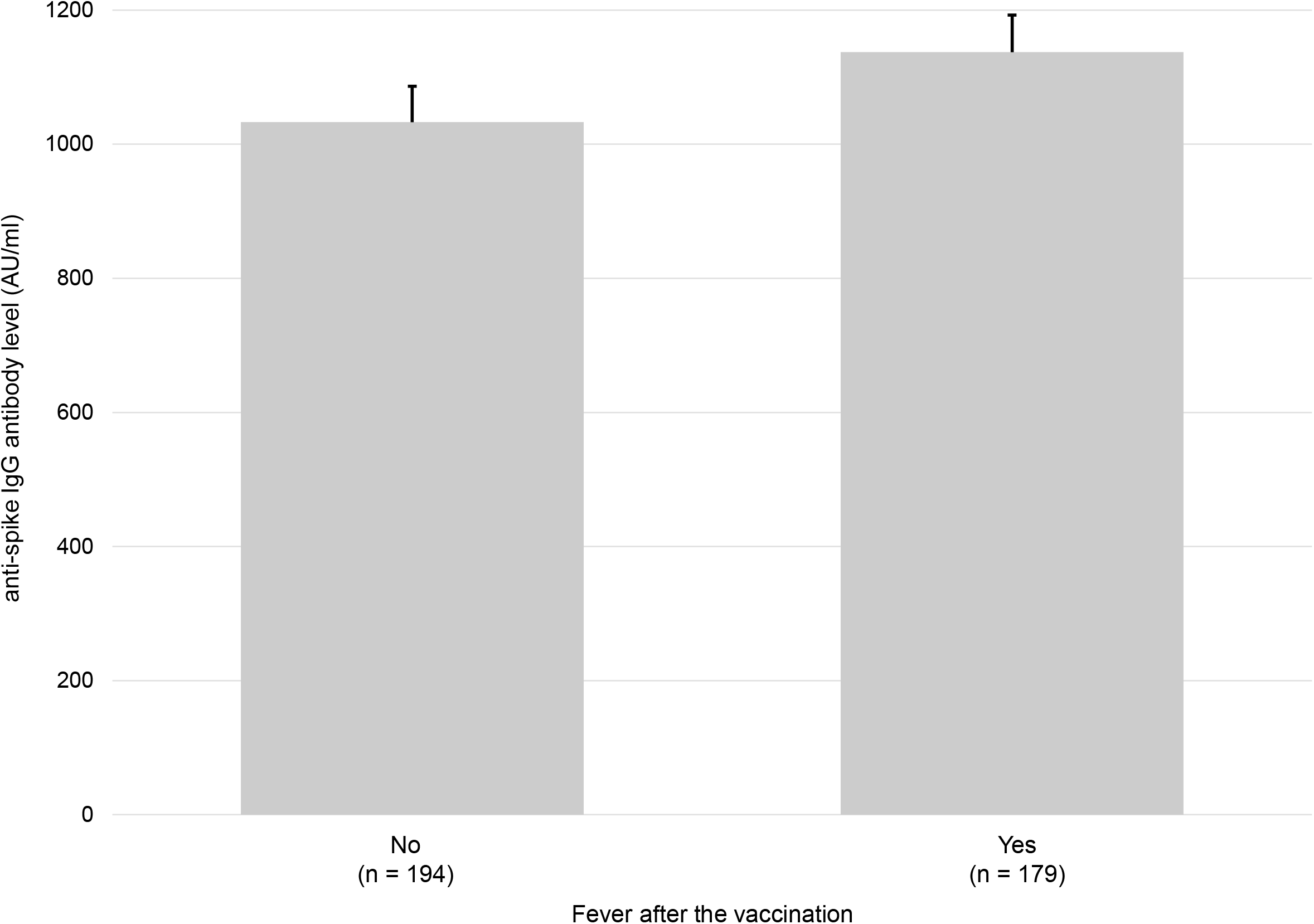
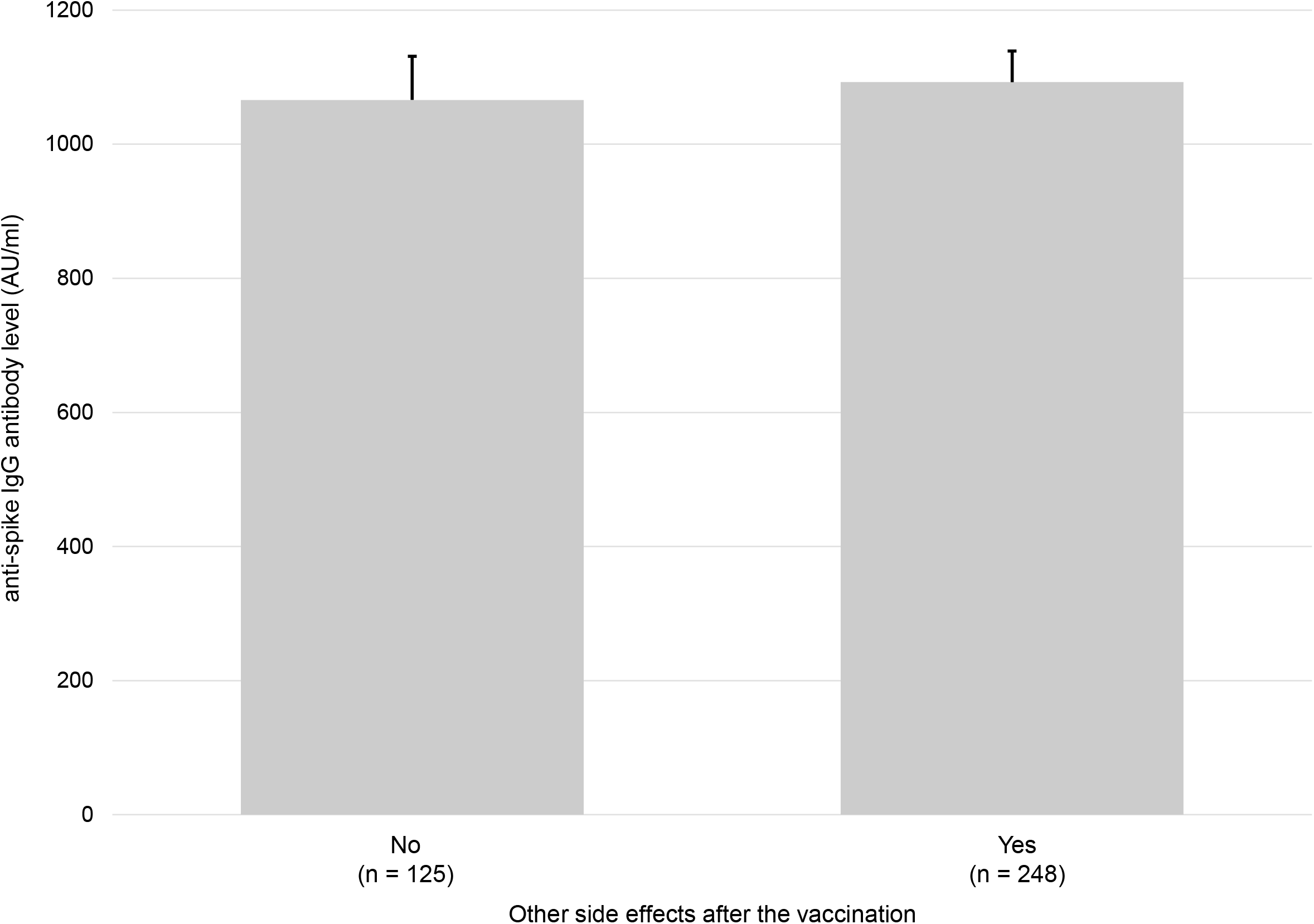
Comparison of anti-spike IgG levels by various groups. The median anti-spike IgG levels were compared by: age group (4a), sex (4b), smoking habits (4c), alcohol drinking habits (4d), allergy (4e), fever after the vaccinations (4f), and other side effects after the vaccinations (4g). The 20s or younger group had a significant higher anti-spike IgG level than any other age group. The 30s group had a significant higher anti-spike IgG level than the 40s and 60s groups. Daily alcohol drinking group had a significant lower anti-spike IgG level than the other groups.

Logistic regression analyses were performed to determine the factors that lead to anti-spike IgG levels above 1000 or 2150 AU/ml at six months after the vaccination (**Figure 5**). Based on the correlation with the plaque reduction neutralization test, an anti-spike IgG level of 1000 AU/ml is considered approximately 60% effective, and that of 2150 AU/ml is considered around 80% effective in preventing infection [17]. In univariate analyses, the factors that significantly influenced anti-spike IgG levels above 1000 AU/ml were age and past smoking history (**Figure 5a**). The factors that significantly influenced anti-spike IgG levels above 2150 AU/ml were age and fever after the vaccination (**Figure 5c**). In multivariate analyses, only age affected anti-spike IgG levels above 1000 AU/ml (**Figure 5b**), while age and alcohol drinking habits affected anti-spike IgG levels above 2150 AU/ml (**Figure 5d**). None of the 53 participants with daily alcohol drinking habits had anti-spike IgG 2150 AU/ml or higher.

**Figure 5.**
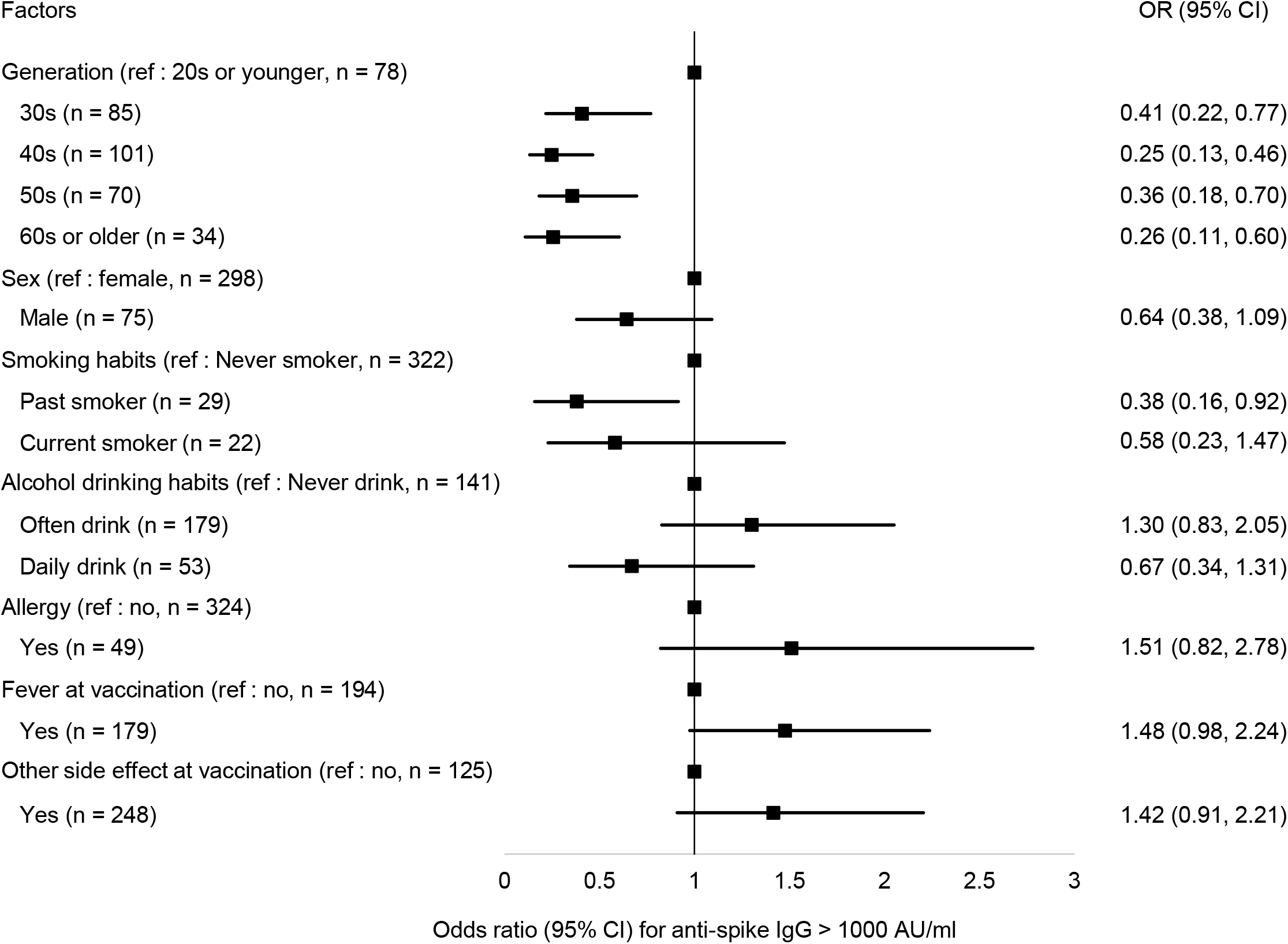
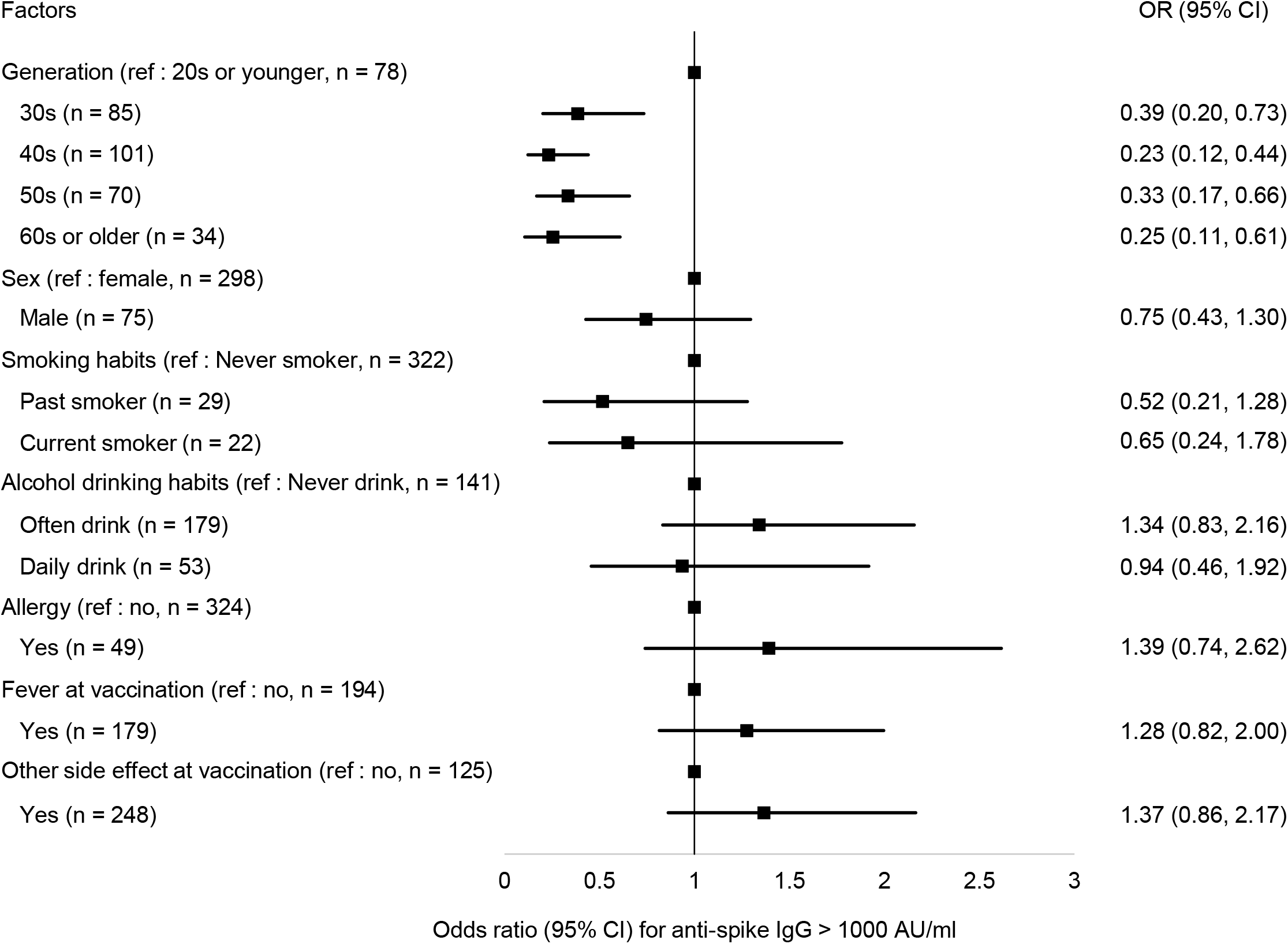
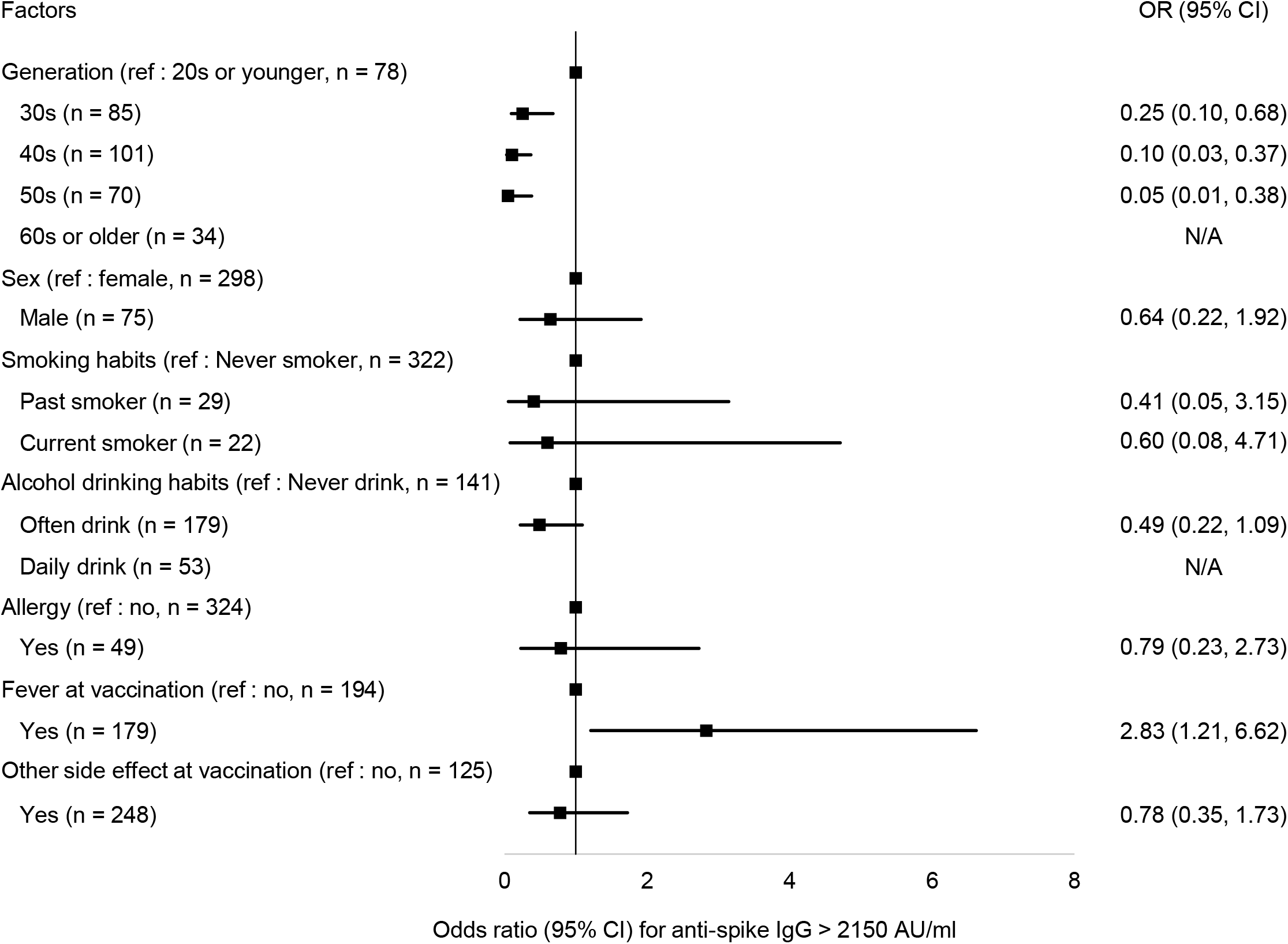
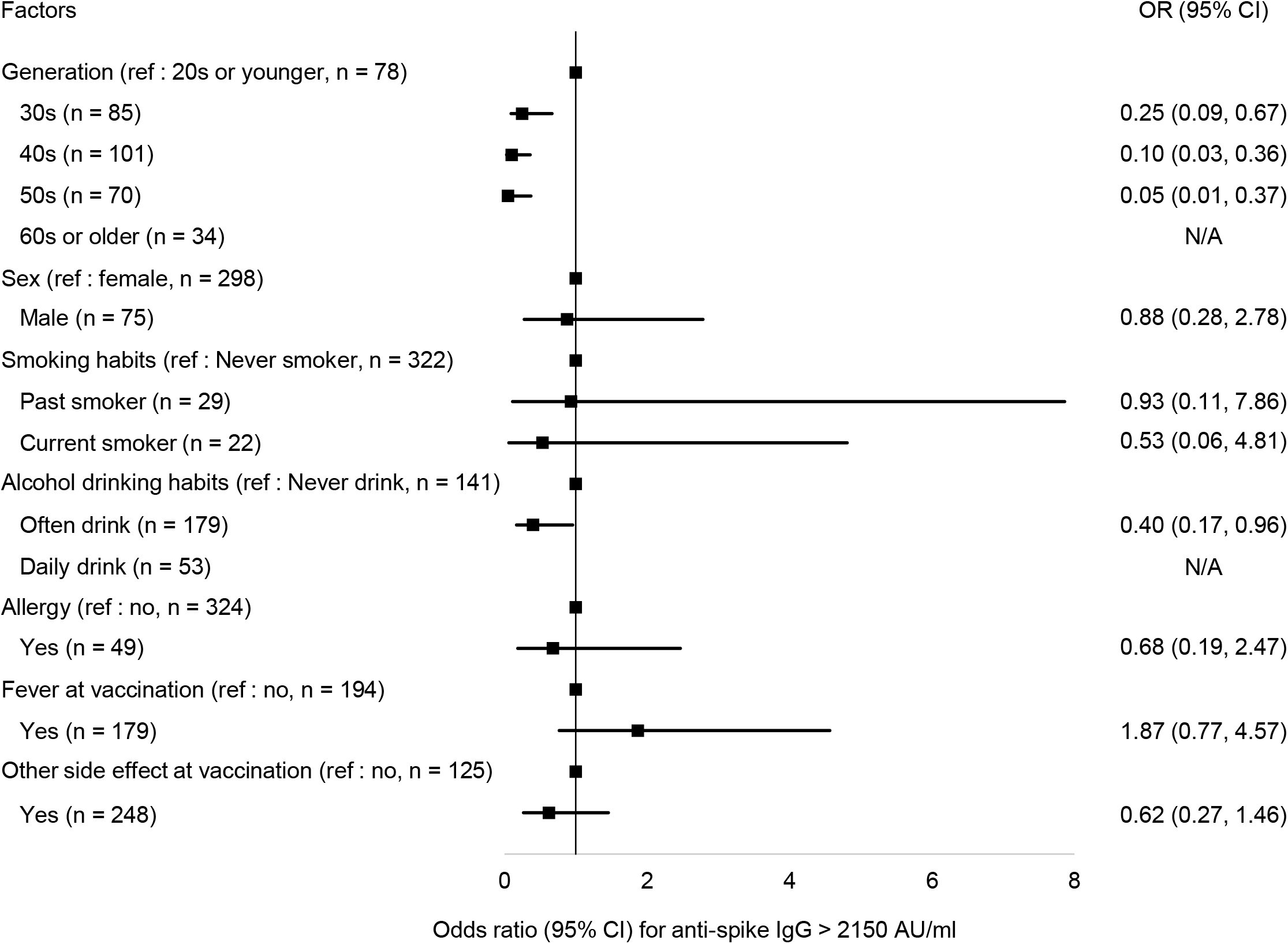
Contributed factor for anti-spike IgG level ≥ 1000 and ≥ 2150 AU/ml. Odds ratios for having ant-spike IgG level ≥ 1000 (5a for univariate and 5b for multivariate) and ≥ 2150 AU/ml (5c for univariate and 5d for multivariate) are shown. Adjusted factors in the multivariate logistic model were sex, generation, smoking habit, drinking habit, allergy status, and the interval days between the second vaccination and anti-spike IgG measurement. In the multivariate analyses, age significantly affected the anti-spike IgG levels ≥ 1000 and ≥ 2150 AU/ml, alcohol drinking habits significantly affected the anti-spike IgG level ≥ 2150 AU/ml.

## Discussion

Vaccination against SARS-CoV-2 has proved to be a highly effective strategy for reducing COVID-19 infection. However, it is still unclear how long the protective effect maintains after the vaccination or what level of anti-spike IgG level is required to protect against COVID-19 infection. Therefore, we conducted one prospective study (Analysis-1) and one cross-sectional study (Analysis-2).

In the prospective study, we found a significant waning of anti-spike IgG level during six months after receiving the second dose of the BNT162b2 vaccine in 49 health care workers. The anti-spike IgG levels at six months after the second vaccination were less than 10% of those at one month after the second vaccination. Previous studies have reported that anti-spike IgG level drops to approximately 10% within six months of vaccination, and our results are consistent with previous studies [18, 19]. A study of 98 healthy subjects reported that in addition to IgG, 50% neutralizing antibodies also dropped to about 20% over six months, suggesting decline efficacies in preventing infection six months after the vaccination [19]. Moreover, we have previously reported that the elderly had less than 30% of anti-spike IgG levels one month after the vaccination compared to subjects under 60 years old [20]. Based on these results, the third vaccination may be desirable for the elderly because the efficacies of the vaccine in preventing infection are considerably reduced six months after the vaccination.

The cross-sectional study found anti-spike IgG levels associated with age and alcohol drinking habits. Compared to the group of 20s or younger, the odds ratios of having an anti-spike IgG level of 1000 or higher six months after the second vaccination were less than 0.5 in other age groups. This trend did not change even after adjustment for sex, smoking habits, alcohol drinking habits, allergies, and days since vaccination. The odds ratios for anti-spike IgG levels of 2150 or higher six months after the second vaccination were similar to that for anti-spike IgG levels of 1000 or higher. Participants with alcohol drinking habits had a significantly lower odds ratio of having an anti-spike IgG levels of 2150 or higher after six months than those without alcohol drinking habits. Especially among those who drank alcohol daily, none of whom had an anti-spike IgG levels of 2150 or higher. These results are also consistent with previous studies [19, 21]. In our previous study, the median anti-spike IgG level of the elderly in the nursing home and the long-term care units was approximately 2000 AU/ml one month after the vaccination [20]. Thus, vaccine efficacy in the elderly may be lower than in the younger.

Some limitations of this study should be noted. We only assessed anti-spike IgG levels, lacking neutralizing antibodies and cellular immunity, which prevent severe disease. The sample size of the whole cohort was small and only 10% of the participants were over 60 years old. In addition, most of the participants did not have any comorbidities. Further extensive studies including participants with various backgrounds and assessing the neutralizing antibodies and cellular immunity are necessary.

## Conclusions

Six months after the vaccination, the anti-spike IgG level was substantially low, especially among persons 40 years of age or older and among persons having alcohol drinking habits.

## Data Availability

All data produced in the present study are available upon reasonable request to the authors.

## Abbreviations

COVID-19: coronavirus disease 2019
IgG: immunoglobulin G
AU/ml: arbitrary units per milliliter
AST: aspartate aminotransferase
ALT: alanine aminotransferase
γ-GTP: γ-glutamyl transpeptidase

